# Development and Clinical Validation of Lightweight, Multimodal Machine Learning Models for Smartphone-Based Cataract Detection and Classification

**DOI:** 10.1101/2025.11.14.25340278

**Authors:** Vishnu Kannan, Jordan Shuff, Arushi Acharya, Rengaraj Venkatesh, Nakul S. Shekhawat, Kunal S. Parikh

## Abstract

Globally, cataract remains the leading cause of blindness, affecting over 100 million people, with a disproportionate burden in low- and middle-income countries where access to ophthalmologists is limited. Although cataract surgery can restore vision almost immediately, timely diagnosis and referral remains a major barrier to care. We developed and prospectively evaluated lightweight, multimodal machine learning models capable of classifying lens status on a smartphone, enabling accessible screening in low-resource environments. We trained and evaluated both early and late fusion model architectures to classify lens status as clear, immature cataract, mature cataract, or pseudophakia using 6,794 anterior segment eye images captured using Scout™ smartphone-based diffuse illumination system paired with clinical data (age, visual acuity, and pinhole acuity) from 2,956 patients from three eye hospitals in India. The early fusion model, which jointly integrates image and clinical features via a learnable gating mechanism, achieved superior performance (AUROC=0.98) compared to late fusion. Model interpretation using feature importance and Grad-CAM revealed that early fusion effectively balanced visual and clinical parameters, mirroring ophthalmologist diagnostic reasoning. Prospective on-device evaluation in 210 patients at the Aravind Eye Hospital demonstrated equivalent performance, achieving an AUROC of 0.96, confirming robustness and real-time feasibility on mobile hardware. These results demonstrate the first prospectively validated, on-device, multimodal cataract machine learning model, demonstrating the feasibility of instant, offline cataract classification and referral in low-resource environments. This advance has potential to broaden cataract screening, allowing minimally trained workers to screen and refer patients, and enabling earlier diagnosis, referral, and treatment in underserved populations.

## 1. Introduction

Cataract formation is associated with increased opacification of the lens of the eye, leading to moderate-to-severe visual impairment (MSVI) or blindness in more than 100 million individuals globally.^1^ The incidence of cataract is increasing as the world’s population ages with a disproportionate burden in low- and middle-income countries (LMICs) where cataracts are a leading cause of total disability adjusted life years due to poverty, lack of access to care, and increased UV exposure.^1,2^ Cataract surgery almost immediately restores vision, and has been shown to significantly improve physical and mental health, income, productivity, and quality of life.^3^ Despite its cost-effectiveness and nationwide campaigns to improve surgical coverage, India accounts for the greatest burden of cataract.^4^ The shortage of ophthalmologists (1:91,000 ophthalmologists:patients) remains a key bottleneck to diagnosis and treatment of underserved populations. In particular, rural populations are more likely to suffer blindness or MSVI due to geographic barriers to eye screening, lack of access to specialists, costs of travel to urban centers for care, and lack of awareness regarding effective treatments for eye diseases.^5,6^

Outreach eye camps have led to reductions in the proportion of cataract blindness and visual impairment; however, they often fail to reach the most remote communities with women, elderly, poor, and rural patients disproportionally affected.^7,8^ Moreover, pen light exams used in these settings still require trained personnel and are limited in capturing early stages of disease, classifying severity, and documenting outcomes.^9^ In contrast, gold standard cataract diagnosis provides quantitative grading of different types of cataract, but requires an experienced ophthalmologist and expensive slit lamp equipment, further limiting accessibility.^10^ There is a significant unmet need for simple, low-cost approaches that de-skill and decentralize cataract screening to reach underserved populations while providing accurate classification of lens status and high triage specificity.^11^

Emerging portable anterior segment eye imaging systems and artificial intelligence (AI)-based diagnostic models have potential to broaden access to cataract screening. Several portable, smartphone-based, handheld slit lamps have been developed to enable cataract screening beyond tertiary care settings; however, they are often expensive, difficult to use, limited by camera resolution, and still require significant expertise and training for interpretation.^12,13^ Artificial intelligence (AI)-based systems have shown promise in alleviating the need for expert interpretation, but they are limited by single-modality inputs, small homogenous datasets, dependence on high-end hardware, and/or lack of prospective on-device validation.^14-18^ Moreover, conventional deep learning or large language models often require consistent internet access or lack compatibility with smartphones available in LMICs.^19^

We address these gaps through development of lightweight, multimodal models that can be deployed on accessible smartphones for offline, real-time lens classification and triage. We collected high quality anterior segment eye images of patient populations from three high volume eye hospitals across India using Scout™, a novel, low-cost, smartphone-based diffuse illumination imaging system. We hypothesized that pairing anterior segment images with patient clinical data (e.g., age, visual acuity, pinhole acuity), mimicking clinical decision-making, would result in high model accuracy for lens classification without slit lamp images or ophthalmologist interpretation.^15,17,18^ We developed and assessed the performance of different multimodal model architectures, deployed models onto accessible smartphones, and evaluated accuracy in a real-world prospective study of 210 patients at the Aravind Eye Hospital (AEH).

## 2. Methods

### 2.1 Data Collection

#### 2.1.1 Data collection and ethics statement

Data were collected from consenting patients at AEH (Pondicherry, Tamil Nadu, India), Dr. Shroff’s Charity Eye Hospital (SCEH; Delhi, India and Mohammadi, Uttar Pradesh, India), and Sadguru Netra Chikitsalaya (SNC; Chitrakoot, Madhya Pradesh, India). Community health workers (CHWs) captured eye images using the Scout™ Anterior Segment Eye Imaging System and recorded patient information, including age, gender, and presenting complaints selected from a standardized list (blurry vision near, blurry vision distance, redness, eye pain or irritation, photophobia, and recent eye trauma). Diagnosis labels for each image were extracted from the electronic health record (EHR) based on an ophthalmologist’s clinical diagnosis of the same patient via slit-lamp examination. Visual acuity and pinhole visual acuity were measured by a refractionist and obtained from the EHR. The study protocol was approved by the Institutional Review Boards of Aravind Eye Hospital, Dr. Shroff’s Charity Eye Hospital, Sadguru Netra Chikitsalaya, and the Johns Hopkins University School of Medicine. Informed consent was obtained by CHWs from all participants prior to image capture.

#### 2.1.2. Smartphone image capture

Images were captured using the Scout™ Anterior Segment Eye Imaging System coupled with a Samsung Galaxy S8 smartphone (SM-G950F, Samsung India Electronics Pvt. Ltd., New Delhi, India). Scout™ incorporates a bi-convex magnifying lens (ThorLabs, Newton, NJ) aligned with the smartphone camera’s optical axis. The lens has a 30 mm focal length and a broadband anti-reflective coating effective from 400 to 1,100 nm. Illumination was provided by two white LEDs (Cree LED, Durham, NC) positioned approximately 15 mm apart, each with a color temperature of 4,100 K and a beam angle of ∼120°. The LEDs were powered through the smartphone’s USB port via an on-the-go connection. A cylindrical silicone scope (Dragon Skin 10 Medium; Smooth-On, Macungie, PA) enclosed the eye to block external light and provide stabilization.

#### 2.1.3 Diagnosis criteria

Diagnoses were made via gold standard slit lamp examination by trained ophthalmologists. “Clear crystalline lens” was defined as a lens without any significant nuclear, cortical, or posterior subcapsular opacity visible on in-person ophthalmologist examination.^20^ “Immature cataract” was defined as mild-to-moderate nuclear, cortical, and/or posterior subcapsular lens opacification that, according to the examining ophthalmologist’s clinical assessment, was associated with mild-to-moderate visual impairment on Snellen visual acuity testing and/or mild-to-moderate reduction in patient-reported visual function. “Mature cataract” was defined as a fully developed lens opacity involving the entire nucleus and cortex, resulting in a white or brunescent appearance. In this case, the lens was typically hard and sclerotic, consistent with advanced nuclear and cortical changes, and associated with severe reduction in Snellen visual acuity and marked impairment in visual function.^21^ “Pseudophakia” or “Posterior Chamber Intraocular Lens (PCIOL)” was defined as the presence of an intraocular lens implant following cataract extraction, with absence of the natural crystalline lens.^22^

#### 2.1.4 Dataset preprocessing and overview

Prior to model development, the dataset underwent a preprocessing and filtering pipeline to ensure data consistency and suitability for machine learning. The dataset included patient-level entries containing image identifiers, age, visual acuity, pinhole acuity, and lens status labels. All rows with missing or incomplete values in these fields were removed. Snellen visual acuity was converted to corresponding decimal equivalents according to established visual acuity conversion conventions (e.g., 6/6 = 1.0, 6/60 = 0.1, Finger Counts = 0.014, Hand Movements = 0.005, Light Perception = 0.0016, No Light Perception = 0.0013).^23^ A derived numeric feature captured the improvement in vision with pinhole correction, calculated as the difference between pinhole and unaided acuity decimals. After preprocessing, each entry consisted of a clean image reference, patient age, normalized acuity values, the derived acuity difference, and the encoded lens status label.^23^

The final dataset (**Table 1**) consisted of 6,794 anterior segment eye images captured via Scout™ and corresponding structured non-image data collected from 2,956 patients. Of these patients, 2,214 had a single visit and 742 had multiple visits. Images were captured across three clinical sites: AEH (2,424 images), SNC (1,433 images), and SCEH (190 images). The dataset consisted of 3,821 images from male patients (56.2%). The mean age of patients was 51.4 years ± 15 years. Lens status was categorized as follows: clear crystalline lens (n=3,324 images), immature cataract (n=2,262 images), mature cataract (n=242 images), and pseudophakia (n=966 images). The average visual acuity was 0.53 ± 0.36, which corresponds most closely to 6/12, and the average pinhole acuity was 0.64 ± 0.35, which corresponds most closely to 6/9.

**Table 1.** Demographic representation and clinical characteristics of the training dataset. The total row summarizes values across all lens status types. Averages are listed as mean ± standard deviation.

|  | **Images**<br>**(N)** | **Patients**<br>**(N)** | **Age** | **Gender**<br>**(% male)** | **Visual**<br>**Acuity** | **Pinhole**<br>**Acuity** |
| --- | --- | --- | --- | --- | --- | --- |
| **Total** | **6,794** | **2,956** | **51.4 ± 15.0** | **56.2%** | **0.53 ± 0.36** | **0.64 ± 0.35** |
| **Clear lens** | 3,324 | 1,375 | 40.8 ± 12.3 | 63.7% | 0.74 ± 0.33 | 0.83 ± 0.29 |
| **Immature**<br>**cataract** | 2,262 | 1,400 | 60.4 ± 8.8 | 50.3% | 0.32 ± 0.26 | 0.45 ± 0.29 |
| **Mature**<br>**cataract** | 242 | 189 | 64.7 ± 9.3 | 47.9% | 0.03 ± 0.08 | 0.03 ± 0.09 |
| **Pseudophakia** | 966 | 738 | 63.5 ± 9.8 | 46.5% | 0.46 ± 0.28 | 0.62 ± 0.31 |

#### 2.1.5 Image preprocessing

Each image was resized to 224 × 224 pixels to match the expected input dimensions of each deep learning model.^24^ Following resizing, the pixel values were normalized and standardized using ImageNet mean and standard deviation normalization parameters to maintain compatibility with pretrained feature extractors. During data loading, each image was transformed into a PyTorch tensor to enable batch-based GPU processing.^25^ The same preprocessing pipeline was applied to all training, validation, and test samples to ensure uniformity and reproducibility across the entire dataset.

### 2.2 Image segmentation

Prior to classification training, all anterior segment images were segmented to remove the periorbital features using a YOLOv5 model (v7.0-226-gdd9e338) implemented with PyTorch 2.1.0.^26^ Image preprocessing and annotation were performed using Roboflow (v1.1.7).^27^ All images were resized to 320 × 320 pixels. The segmentation training dataset consisted of 717 images with eyes manually annotated using polygon segmentations of the globe. The model was trained for 100 epochs to optimize detection and segmentation performance.

### 2.3 Model architecture

Three lightweight convolutional neural network (CNN) backbones, ResNet-18, EfficientNet-B0, and MobileNetV3-small, were trained and compared using pre-trained ImageNet weights. These models were selected because of their accuracy and lightweight nature, making them suitable for real-time deployment on smartphones.^24^ For the structured, non-image data consisting of age, visual acuity, pinhole visual acuity, and derived acuity difference, five common machine learning classifiers were evaluated: logistic regression, k-nearest neighbors, decision tree, random forest, and gradient-boosted decision trees (GBDT).^28^ Each model was trained with standardized features and evaluated using cross-validation. The best performing classifier was used for integration into the multimodal late fusion architecture.

In the late fusion design, each modality produced its own classification probabilities: one from the best performing image CNN and one from the best performing tabular GBDT. The two models form an ensemble classifier, and their predictions were combined through stacking. The probability vectors were concatenated and passed to a meta-classifier that learned on the combined probability vector. A GBDT classifier was selected as the meta-classifier because it performs well on small, low-dimensional input spaces and can best form consensus among predictions.^29^ This approach allows the system to leverage each modality independently and integrate them in a data-driven manner. The early fusion network combined visual and tabular information before classification. The selected CNN backbone performs feature extraction to produce a compact feature representation of the input image, and a small multilayer perceptron (MLP) processed the tabular data through fully connected layers with ReLU activations, batch normalization, and dropout. The outputs of the two branches were concatenated, and a learnable gating mechanism adaptively weighted the image and tabular embeddings for each patient before final classification. This structure allows the model to jointly learn how visual and clinical cues interact, while remaining lightweight enough for smartphone deployment. Both model frameworks are shown in **Figure 1**.

**Figure 1.**
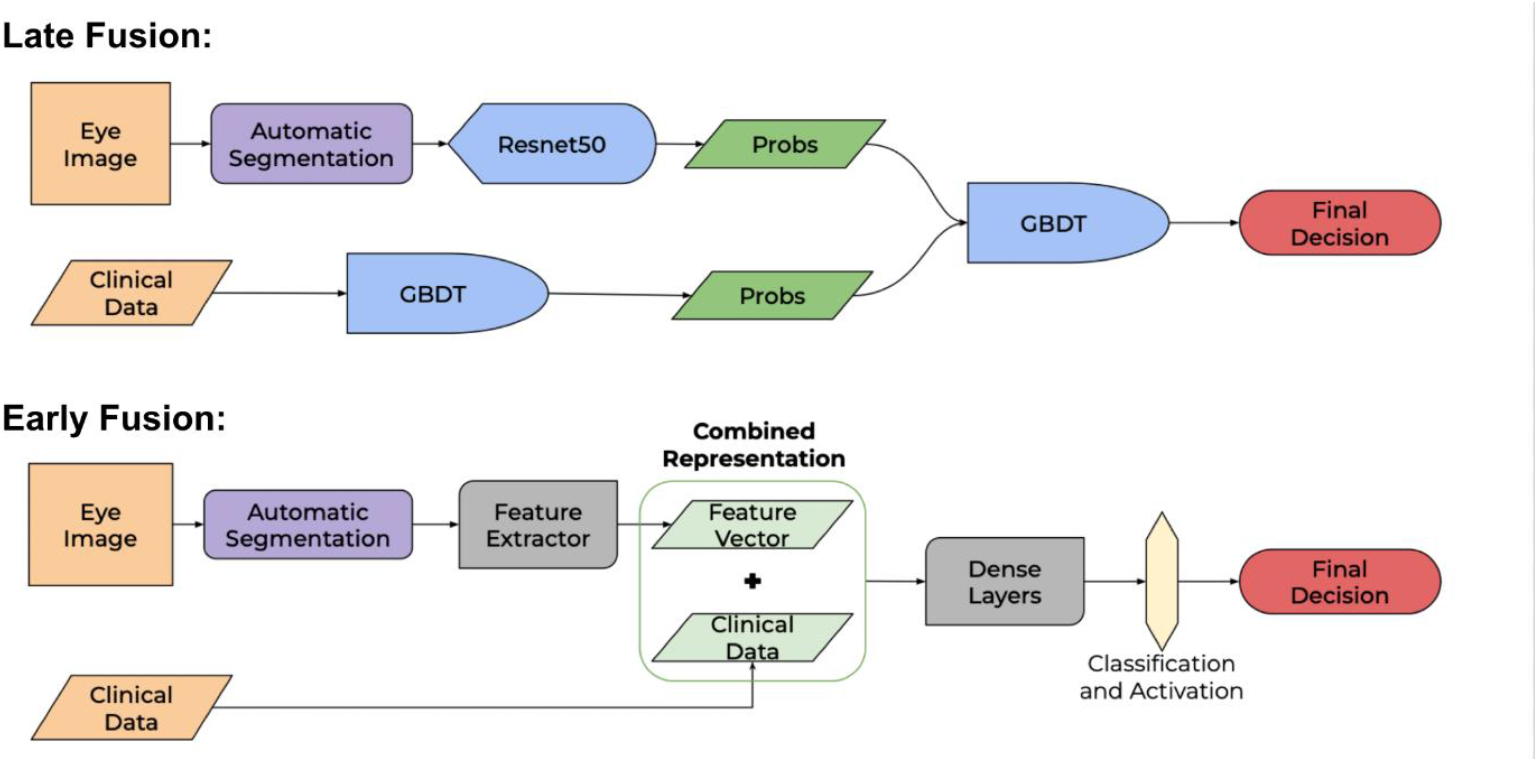
Multimodal model architectures for cataract classification. Schematics of both multimodal machine learning frameworks developed for cataract detection and grading. Late fusion produces a probability vector based on image and clinical data separately and then integrates them for a final prediction. Early fusion concatenates the clinical data with a feature vector and learns from the combined representation. Both architectures use a lightweight CNN backbone for image-based feature extraction, enabling efficient deployment on accessible smartphones.

### 2.4 Training and evaluation

The dataset was split into training, validation, and test sets (64%, 16%, and 20% respectively) using stratified sampling to preserve class balance. The same partitioning was applied consistently across all models to ensure comparability. Image models were trained using cross-entropy loss and class-balanced weights.^30^ Training was performed for up to 30 epochs with early stopping based on validation area under the receiver operating characteristic curve (AUROC), using a patience of five epochs to prevent overfitting.^31^ For the late fusion framework, each base model was trained separately, and out-of-fold predictions from three-fold cross-validation were used to train the meta-classifier. During testing, the predicted probabilities from the image and tabular models were concatenated and passed through the meta-classifier to obtain the final prediction. The model output represented one of four diagnostic classes corresponding to lens status.

Model performance was primarily evaluated using the one-vs-rest area AUROC on the held-out test set. Secondary metrics were assessed, including overall accuracy, and macro-averaged F1-score. 95% confidence intervals were computed using non-parametric bootstrapping with 300 resamples.^32^ Differences between the early and late fusion models were tested using paired bootstrap analyses for classification agreements. ROC curves were generated for each class and for the overall micro-average to visualize discriminative performance. All training, validation, and evaluation procedures were conducted using a fixed random seed to ensure full reproducibility.

### 2.5 Explainability and feature attribution

Feature importance analyses were performed to interpret the contribution of individual clinical variables and modalities. For the tabular data and late-fusion meta-classifier, feature importances were derived directly from the tree-based gain values.^33^ The resulting values were plotted to visualize relative importance. In the late fusion model, this approach also quantified the relative influence of the image-derived probabilities versus tabular probabilities. Linear bias and gate weights obtained from the early fusion model were also examined to gauge relative importance of image and tabular features in classification.^34^

To interpret visual reasoning, gradient-weighted class activation mapping (Grad-CAM) was applied to the image branch of the early fusion model.^35^ Grad-CAM heatmaps were generated from the final convolutional layer and overlaid on the input images to identify regions that contributed most strongly to each classification. To ensure stable and interpretable maps, Grad-CAM was normalized for each image. Maps were computed while the corresponding tabular features were active in the forward pass, ensuring that the heatmap reflected the influence of visual information conditioned on the patient’s clinical context. Visual explanations were generated for four randomly selected data points from each class: two correctly classified, and two incorrectly classified. They were annotated with the true and predicted classes, as well as the prediction confidence.

### 2.6 Prospective validation study

The best-performing model from prior experiments (early fusion model incorporating a ResNet18 backbone) was prospectively validated to assess its diagnostic performance and feasibility in a real-world clinical setting. Following image capture, diagnostic inference was performed directly on the smartphone to determine the model’s capability for fully offline, on-device operation.

#### 2.6.1. Mobile deployment

The model was deployed on a Samsung Galaxy S8 smartphone. Model weights were exported from PyTorch (Torch 2.7.1) to ONNX (v1.17.0) format and subsequently converted to TensorFlow Lite using the onnx2tf framework (v1.28.1). All computations were performed with 32-bit floating-point precision. Input images were resized to 224 × 224 pixels and normalized using ImageNet parameters. The deployed model accepted two inputs: a preprocessed ocular image and a four-element vector of tabular clinical variables. A softmax activation was applied to the output layer to generate class probability scores. The TensorFlow Lite runtime (v2.16.1) executed inference locally on the device without reliance on network connectivity.

#### 2.6.2. Dataset and patient demographics

The validation dataset included 210 anterior segment images from 118 patients. Images were captured by a CHW at AEH using Scout™. Each Patients were recruited from the general ophthalmology and contact lens clinics to ensure sufficient representation of eyes with clear crystalline lenses and achieve a more balanced class distribution. Visual and pinhole visual acuity measurements were determined by a trained refractionist. Diagnostic labels were extracted from the EHR based on gold standard ophthalmologist slit-lamp examinations. Ophthalmologists were masked to model outputs during patient examination. Each eye was assessed individually. The dataset included 80 images from male patients (37.9%). The mean patient age was 55.5 ± 16.5 years. Of the 210 images, 50 were classified as having a clear crystalline lens, 49 as immature cataract, 58 as mature cataract, and 53 as pseudophakia. The mean visual acuity was 0.35 ± 0.36 (approximately 6/18 Snellen equivalent), and the mean pinhole acuity was 0.64 ± 0.35 (approximately 6/9 Snellen equivalent).

## 3. Results

We developed and validated multimodal machine learning models that classify lens status (e.g., clear crystalline lens, immature cataract, mature cataract, pseudophakia) by combining anterior segment eye images with patient clinical data. We hypothesized that a multimodal approach would outperform single mode classification and allow for real-time cataract screening when deployed on edge devices. We developed models to classify based on images and clinical variables individually, compared multimodal late fusion and early fusion strategies, and analyzed interpretability through feature importance and visual attention analysis methods. Lastly, we deployed the best performing model on a smartphone paired with Scout™ for prospective evaluation in a real-world setting by a CHW.

### 3.1 Segmentation results

We applied segmentation to remove periorbital features from diffuse illumination eye images. On a test set of 203 images, the YOLOv5 model trained on images from our dataset achieved excellent precision of 0.989, recall of 0.985, mean average precision at 50% intersection-over-union (mAP50) of 0.982, and a mean average precision across intersection-over-union thresholds (mAP) of 0.896. This trained model was applied to segment all images in the dataset for subsequent classification training.

### 3.2 Dataset and evaluation overview

We analyzed a large, real-world dataset of Scout™ anterior segment eye images paired with minimal, clinically salient structured variables (age, unaided visual acuity, pinhole visual acuity, and a derived acuity difference feature). Models were trained and evaluated on stratified train/validation/test partitions; the primary endpoint was macro one-vs-rest AUROC on the held-out test set with 95% bootstrap confidence intervals. Secondary endpoints included overall accuracy, balanced accuracy, and macro-F1. This evaluation framework reflects current best practice for multi-class clinical AI classification problems and allows class-wise interpretability via confusion matrices and Grad-CAM overlays.

To establish a baseline for image-based classification, we evaluated three lightweight CNN architectures trained only on segmented eye images. We selected ResNet-18, EfficientNet-B0, and MobileNetV3-small based on their lightweight nature and ease for smartphone deployment.^24^ Each model was initialized with ImageNet pretrained weights and fine-tuned on our dataset using the 64% training, 15% validation, 20% test splits. Performance was primarily assessed using macro one-vs-rest AUROC, with accuracy, balanced accuracy, and macro-F1 score as secondary metrics (**Table 2**).

**Table 2.** Performance of lightweight CNN architectures trained on Scout™ images. All three CNN architectures showed similar high performance, with ResNet achieving the highest AUROC.

| Model | AUROC | Accuracy | Balanced Accuracy | Macro F1 |
| --- | --- | --- | --- | --- |
| Resnet-18 | 0.969 | 0.872 | 0.888 | 0.885 |
| EfficientNet-B0 | 0.968 | 0.868 | 0.859 | 0.868 |
| MobileNetV3-small | 0.967 | 0.837 | 0.868 | 0.832 |

All three lightweight CNN architectures had similar performance with AUROC values near 0.97. Resnet-18 achieved the highest performance (AUROC=0.969), followed closely by EfficientNet-B0 and MobileNetV2-Small. The consistent high and narrow range of performance suggests that the visual features captured by Scout™ are sufficiently informative to support accurate distinction of lens status regardless of CNN design. Subsequent multimodal model development was performed using ResNet-18 as the image backbone due to its superior performance.

To understand the overall and relative value of clinical variables in discrimination, we evaluated five machine learning classifiers using only age, visual acuity, pinhole acuity, and the calculated difference between pinhole and visual acuity (**Table 3**). This assessment was also used to understand the diagnostic information that could be extracted from the clinical data and determine which classifier would be most suitable for integration in a multimodal framework.

**Table 3.**
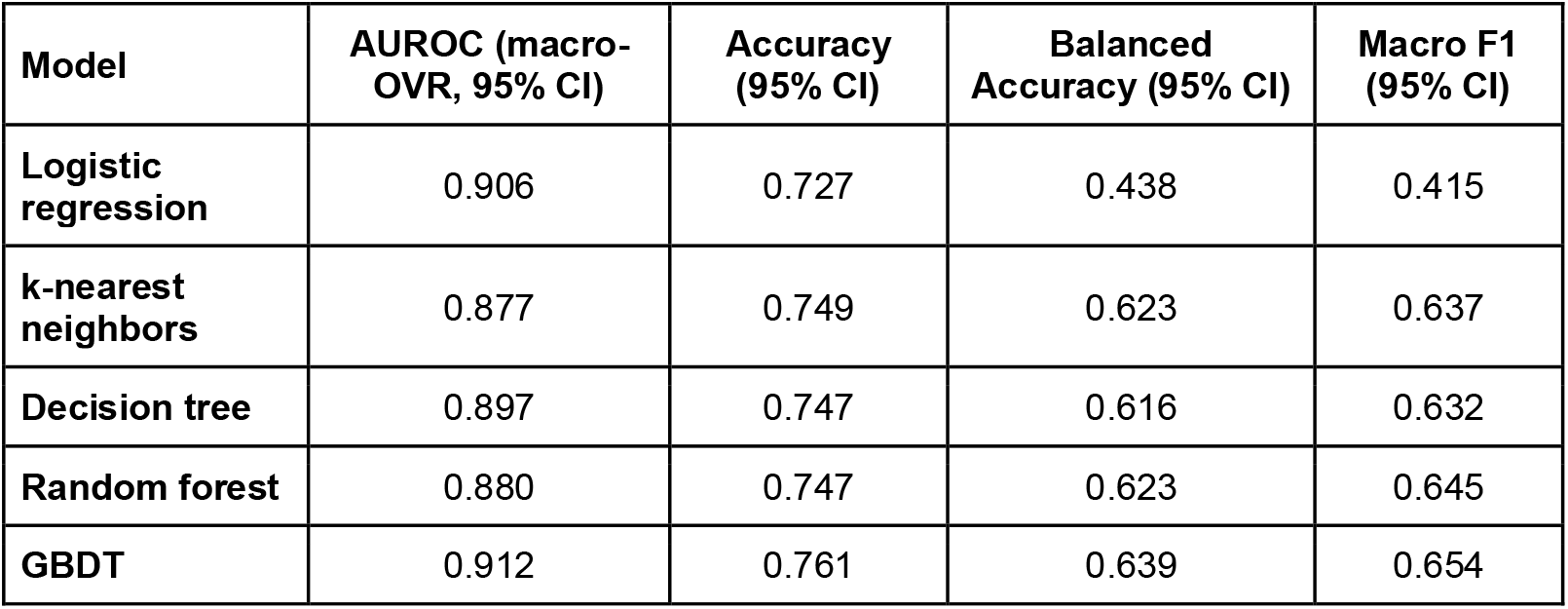
Performance comparison of tabular data classifiers using only clinical variables. GBDT achieved the highest AUROC and accuracy, capturing relationships between clinical features and lens status. The performance across all classifiers indicates that these clinical variables carry substantial diagnostic value.

GBDT achieved superior performance among these classifiers with an AUROC of 0.91 and the highest accuracy of 0.76. The logistic regression model resulted in the second highest AUROC (0.91), but the lowest accuracy (0.73) with particularly low balanced accuracy (0.44). The ensemble methods, random forest and GBDT, demonstrated more balanced performance. All classifiers achieved AUROC above 0.87, indicating that the clinical variables alone carry substantial information predictive of lens status. However, lower balanced accuracy and F1-scores suggest it has limitations in distinguishing certain types of lens status, and that more information may be obtained when combined with image data context.

Age had the greatest influence on GBDT classification by a significant margin, with over 70% of the predictive weight (**Figure 2**). This reflects the strong association between age and cataract severity, particularly in settings where there is limited access to eye care.^36^ Visual acuity and pinhole acuity had almost identical importance in classification. This suggests that the model may be using both metrics to gauge severity while distinguishing vision loss due to cataracts from other causes. The acuity difference contributed minimally despite established clinical utility. This suggests that the absolute visual and pinhole acuity functionally capture the correctability information that the difference was intended to encode. Overall, these results show that the GBDT classifier learns a clinically interpretable feature hierarchy while achieving relatively high performance.

**Figure 2.**
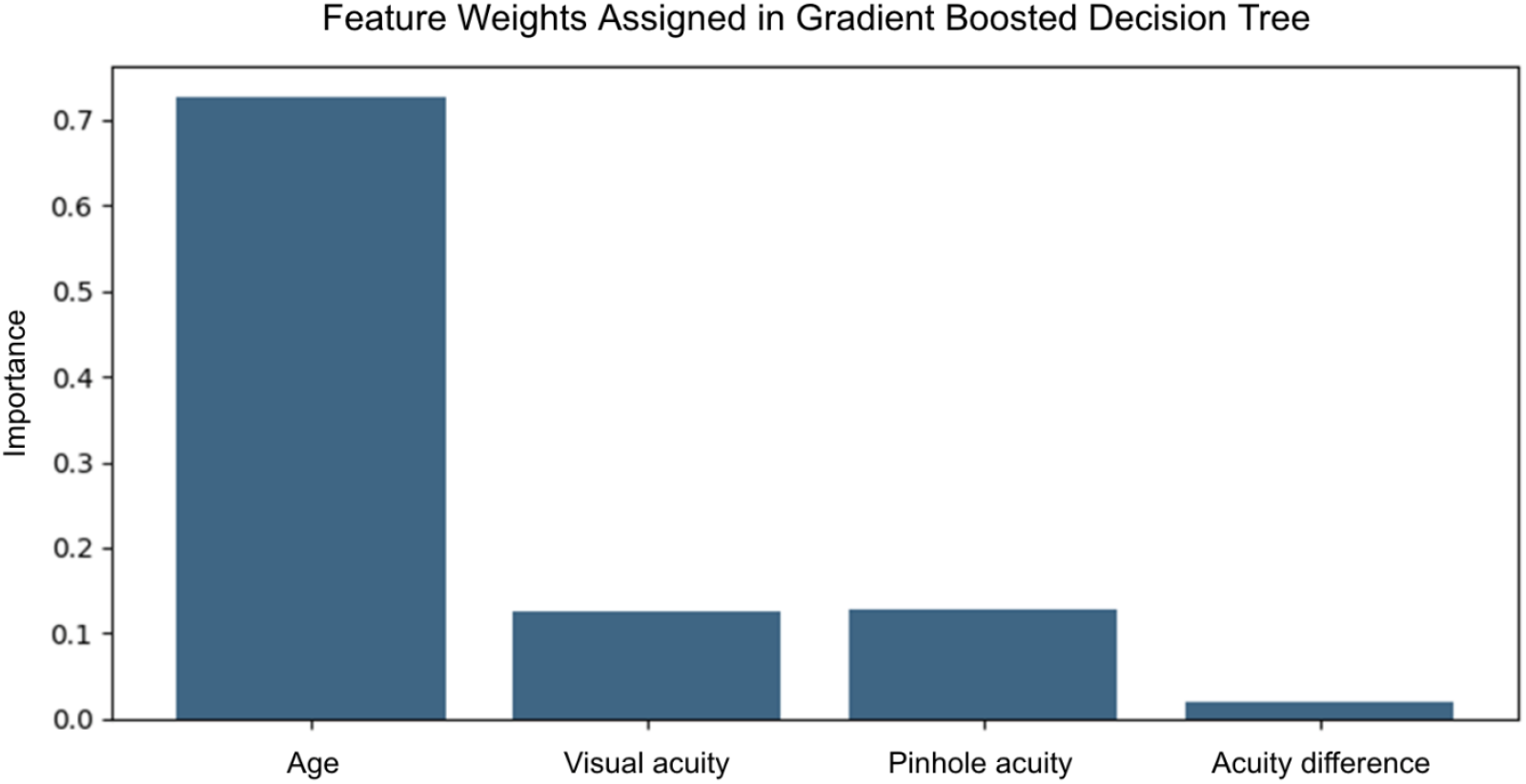
Feature importances for clinical data classification. Relative importance of each clinical variable in the GBDT classifier trained on age, visual acuity, pinhole visual acuity, and their difference. Age was the dominant predictor of lens status, followed by visual and pinhole acuities, which contributed nearly equally.

We next investigated whether integrating the image and tabular data modalities could improve classification accuracy. We compared two fusion strategies: late fusion, which combines independent predictions from the trained image and tabular data models using a meta-classifier, and early fusion, which learns based on a joint representation of the modalities through a network with adaptive gating. We hypothesized that early fusion would enable the model to capture complex interactions between visual and clinical features, more closely mimicking interpretable decision-making processes used by clinicians.

Early fusion achieved superior performance with an AUROC of 0.98, representing a small improvement over late fusion at 0.97 (**Table 4**). The overall high AUROC suggests that early fusion distinguishes lens status classes by learning adaptive relationships between visual and clinical features. Surprisingly, the performance difference between models was small: late fusion demonstrated marginally higher overall accuracy (0.89 vs 0.87) and macro-F1 score (0.88 vs 0.86) in comparison to early fusion. The exceptional performance of both models may indicate that we are reaching an upper limit for lightweight model performance. However, because AUROC measures performance across all thresholds and accuracy is affected by class distributions, we hypothesized that early fusion may perform better in a real-world setting. Moreover, early fusion architecture’s learnable gating mechanism enables dynamic weighting of image and tabular embeddings on a per-sample basis, which may allow the model to rely more heavily on clinical data when images are degraded or ambiguous, and vice versa. Thus, we selected the early fusion model for prospective validation and mobile deployment.

**Table 4.** Performance comparison of multimodal fusion strategies. Early fusion achieved the highest AUROC although balanced accuracy and macro-F1 scores were nearly equivalent between both approaches, suggesting similar performance on minority classes.

| Model | AUROC<br>(macro-OVR,<br>95% CI) | Accuracy<br>(95% CI) | Balanced<br>Accuracy<br>(95% CI) | Macro F1<br>(95% CI) |
| --- | --- | --- | --- | --- |
| Late Fusion | 0.973 | 0.889 | 0.882 | 0.880 |
| Early Fusion | 0.979 | 0.874 | 0.883 | 0.859 |

Confusion matrixes were further examined to study the distribution of predictions across lens status categories for both architectures. This provides a better understanding of classification patterns and error nodes to compare the models. The confusion matrices reveal distinct classification patterns between early and late fusion (**Figure 3**). Both models performed nearly perfectly on PCIOL. Early fusion correctly classified slightly more mature cataract cases (43 vs 39), suggesting small improvement when features are considered jointly. However, early fusion also misclassified more clear lens eyes as immature cataracts (62 vs 45). This pattern may reflect borderline cases where early lens changes are visible on examination but have not yet significantly impacted visual function. Thus, the model’s bias towards disease detection could lead to over-referral but would not miss actionable pathology. Late fusion demonstrated more balanced performance in distinguishing clear lens from immature cataract, possibly because the meta-classifier can independently weight the high-confidence clear lens predictions from clinical data against the comparatively ambiguous image features. Both fusion strategies showed the highest error rates for mature cataract classification. This pattern likely reflects the relative scarcity of mature cataract examples in the training dataset (3.56% of the training dataset), limiting the models’ exposure to the full range of dense lens opacities and resulting in occasional confusion with immature cataract. These results demonstrate that fusion strategy selection involves trade-offs in sensitivity and specificity across different lens status categories.

**Figure 3.**
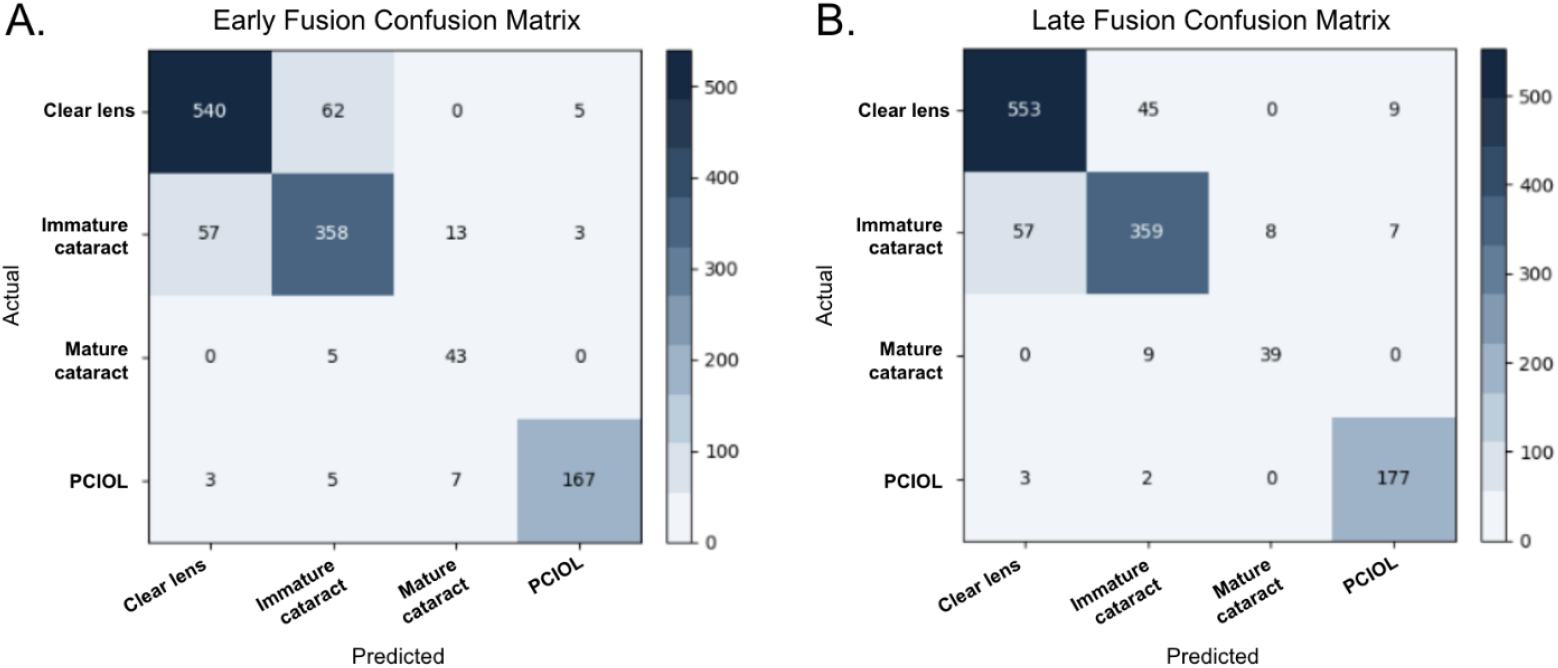
Confusion matrices for early and late fusion models. Confusion matrices for **(A)** early and **(B)** late fusion frameworks. Darker diagonal cells indicate correct classifications. Both models achieved near perfect classification of pseudophakia (PCIOL), which reflects its distinctive appearance and visual function. Early fusion classifies mature cataract more accurately, but a higher rate of false positive immature cataract predictions for clear lens. Both models showed the highest error rates for mature cataract classification.

Feature importance patterns were analyzed based on the late fusion meta-classifier and gating weights in the early fusion model. Gating weights from each sample were averaged per class and used for analysis. These patterns provide insight into the decision-making patterns in each architecture (**Figure 4**). The late fusion classifier demonstrated hierarchy in feature use. The tabular model’s probability for clear lens received the highest weight, indicating that clinical variables are more reliable in identifying healthy eyes compared to image data. The image models’ probabilities for immature cataract and pseudophakia received high weights as well, indicating that the model relies more on visual features for distinguishing these classes. This also suggests that clinical variables could be more valuable for ruling out pathology, while image features help classify disease subtypes. The early fusion model’s gating mechanism revealed more balanced modality weighting with average weights of 0.81 for image features and 0.91 for tabular features. This suggests that early fusion learns to dynamically integrate both modalities rather than heavily prioritizing one over the other. However, class-specific analysis revealed some variation: for clear crystalline lens classification, the gate weights favored tabular features more strongly, proving the diagnostic value of normal age and acuity. For other lens status categories, the weights converged toward unity, indicating more symmetric reliance on both modalities. The model’s linear bias toward tabular features was minimal (-0.0301 for image, +0.0299 for tabular), further confirming balanced baseline integration. This adaptive gating behavior may enable the model to compensate for degraded or ambiguous inputs in either modality, for example, relying more heavily on clinical data when images are affected by motion blur, poor illumination, or patient non-compliance, and vice versa, when visual function measurements are unreliable.

**Figure 4.**
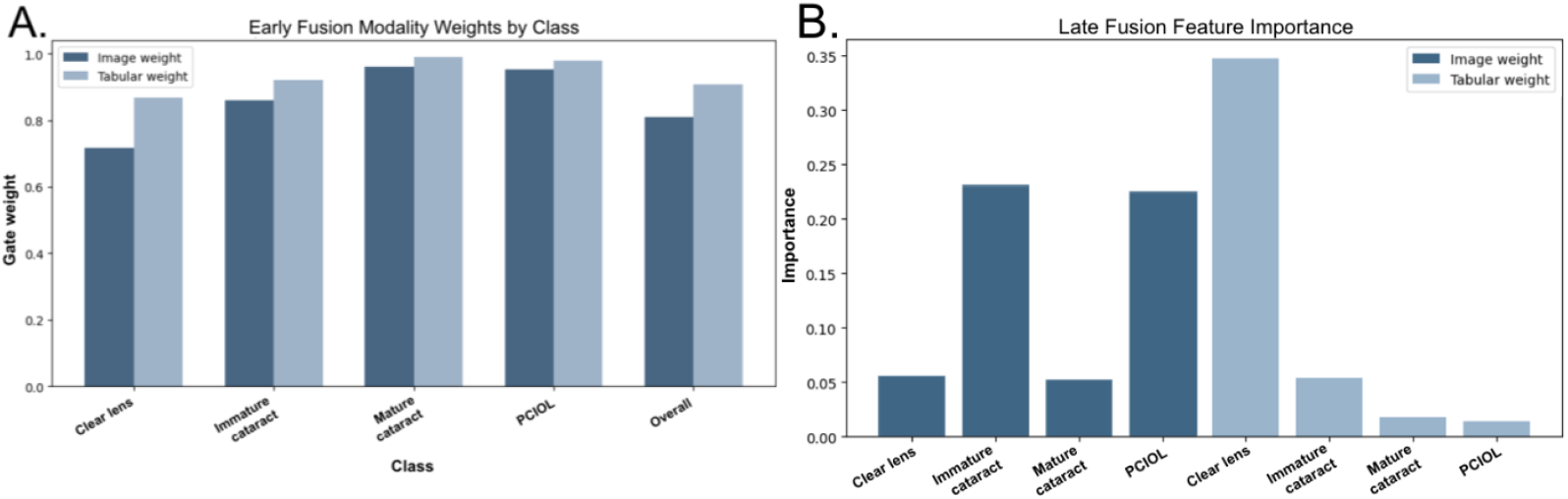
Late fusion and early fusion feature importance analysis. **(A)** Early fusion gate weights per class and **(B)** late-fusion meta-GBDT feature importances. The early fusion model shows slightly higher weight for the tabular data overall. The largest discrepancy is in clear lens, with more clear weighting for the tabular over image data. For clear lens, the late fusion model has a strong preference for the tabular probability, which took almost 35% of total weight. In contrast, the image model was favored for immature cataract and PCIOL.

To understand the interpretability of the image component of the early fusion model, we applied Grad-CAM to generate maps that highlight areas that most strongly influenced classification decisions. A random selection of two correct and two incorrect samples were chosen from each class to analyze (**Figure 5**). This assessment reveals whether the model has learned clinically meaningful visual features from image data. Grad-CAM analysis showed that the early fusion model consistently highlights clinically relevant regions across lens status categories. Attention is tightly concentrated on the lens and pupil in correctly classified cases, with minimal activation on other regions, even in images with poor centration or off-axis patient gaze, demonstrating robust feature identification. This proves interpretability for the early fusion model. Incorrectly classified images were more likely to show more dispersed attention. Correctly classified images also tended to have higher confidence predictions, which may be connected to the more sharply defined heatmaps. High interpretability could facilitate better performance for healthcare professionals, regardless of whether images are classified correctly.^37^

**Figure 5.**
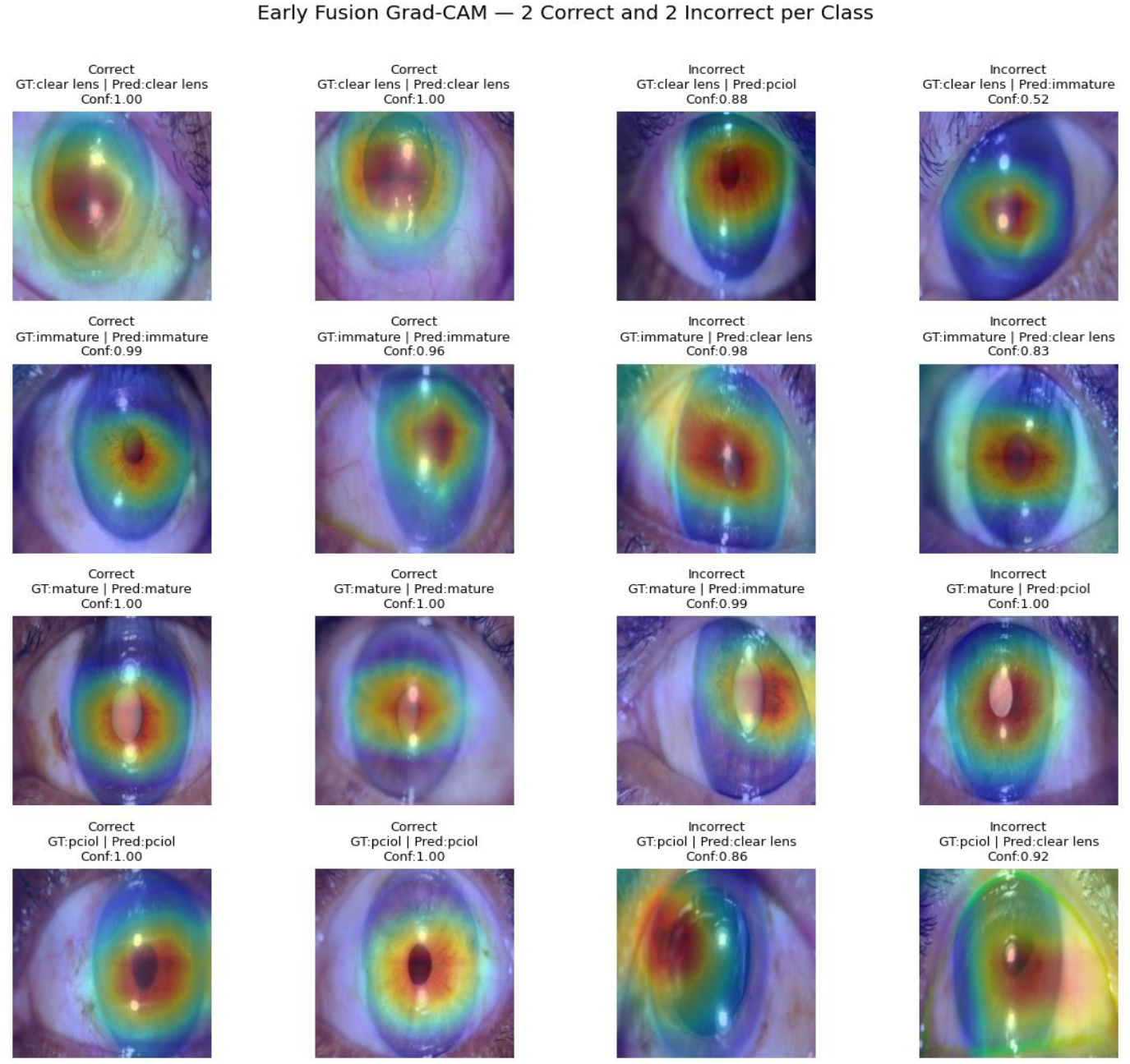
Grad-CAM visualization and confidence scores for the early fusion model. Representative examples showing Grad-CAM overlays for early fusion model for two randomly selected correct (left) and incorrect (right) predictions in each class. Red regions indicate areas of high attention, while blue indicates low attention. The model consistently focuses on the lens and pupil, which are the relevant features for lens status assessment, suggesting clinical interpretability.

### 3.3 Prospective evaluation of baseline and fusion models

We performed a prospective study of the early fusion multimodal model to evaluate performance in a real-world setting following mobile model deployment for offline use. In this study, a trained CHW at AEH used Scout™ to capture 210 anterior segment images of patients with clear lens, immature cataract, mature cataract, or pseudophakia (**Table 5**). Model outputs were compared to gold standard in-person ophthalmologist examination via slit lamp. Both CHWs and ophthalmologists were masked to each other’s diagnoses.

**Table 5.** Demographic and clinical characteristics of prospective study patients, stratified by lens status. Averages are listed as mean ± standard deviation.

|  | **N**<br>**images** | **Age** | **Gender**<br>**(% male)** | **Visual**<br>**Acuity** | **Pinhole**<br>**Acuity** |
| --- | --- | --- | --- | --- | --- |
| **Total** | **210** | **55.5 ± 17** | **37.9%**<br>**(n=80)** | **0.35 ± 0.4** | **0.47 ± 0.39** |
| **Clear lens** | 50 | 29.8 ± 7.6 | 50.0%<br>(n=25) | 0.76 ± 0.4 | 0.93 ± 0.17 |
| **Immature**<br>**cataract** | 49 | 61.8 ± 8.9 | 38.8%<br>(n=19) | 0.23 ± 0.2 | 0.36 ± 0.28 |
| **Mature**<br>**cataract** | 58 | 64.1 ± 9.5 | 36.2%<br>(n=21) | 0.02 ± 0.1 | 0.02 ± 0.05 |
| **Pseudophakia** | 53 | 63.6 ± 8.9 | 28.3%<br>(n=15) | 0.48 ± 0.3 | 0.67 ± 0.21 |

The multimodal early fusion ResNet18 model demonstrated exceptional diagnostic performance across all lens categories (**Table 6**). The model achieved an accuracy of 0.98 for clear crystalline lens, 0.91 for immature cataract, 0.95 for mature cataract, and 0.98 for pseudophakia. The overall AUROC (macro) was 0.96 (**Figure 6**). The average execution time per image on the smartphone was 313.8 ± 2.4 ms, demonstrating the feasibility of near real-time, on-device inference. Overall, the results indicate that the early fusion model can accurately classify lens status across all categories while operating efficiently on mobile hardware in a real-world clinical setting.

**Table 6.** Performance of the early fusion model deployed on a smartphone in a prospective patient study.

|  | **Precision** | **Recall** | **Accuracy**<br>**(95% CI)** | **Balanced**<br>**Accuracy**<br>**(95% CI)** | **Macro F1**<br>**(95% CI)** |
| --- | --- | --- | --- | --- | --- |
| **Clear lens** | 0.942 | 0.980 | 0.981 | 0.981 | 0.961 |
| **Immature**<br>**cataract** | 0.843 | 0.878 | 0.933 | 0.914 | 0.860 |
| **Mature**<br>**cataract** | 0.946 | 0.914 | 0.962 | 0.947 | 0.930 |
| **Pseudophakia** | 1.000 | 0.962 | 0.990 | 0.981 | 0.981 |

**Figure 6.**
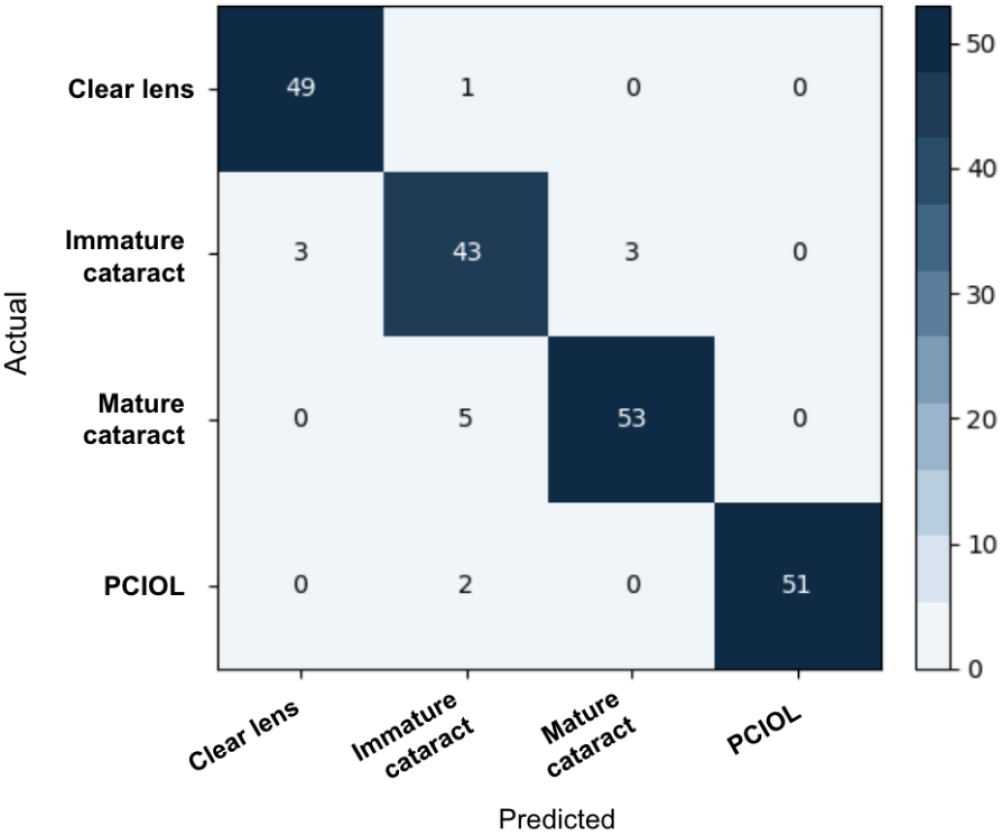
Confusion matrix of early-fusion model deployed on a smartphone in prospective validation study. The model achieves high accuracy across lens status categories, the highest being for pseudophakia (0.99) and clear lens (0.98). The overall AUROC was 0.956 which represents only a minor reduction in performance from in silico testing (0.979), confirming robust real-world deployment.

## 4. Discussion

Cataract remains the leading cause of avoidable blindness globally, with a disproportionate burden in LMICs where there is also a shortage of trained eye care providers. Conventional outreach approaches, including vision centers and eye camps, have helped expand access to eye screening and care, but require significant investment and have limited scalability, leading to lack of access among women, older adults, and rural patients. A lightweight, smartphone-based, offline model with low latency has potential to enable task shifting to CHWs or providers in primary care settings, broadening access to eye screening and facilitating timely referral for treatment. Importantly, in settings where access to eye care is limited, other vision-threatening eye conditions, such as glaucoma, are often only diagnosed following referral for cataract.^40^ Thus, accessible cataract screening has potential to significantly reduce avoidable blindness while also promoting overall eye health.

We developed, validated, and deployed a lightweight, multimodal model that grades lens status from smartphone-based diffuse illumination images enhanced with clinical variables (age, visual acuity, pinhole acuity, and the difference between visual and pinhole acuity). Early fusion was the best performing model achieving 0.98 AUROC for classification between clear lens, immature cataract, mature cataract, and pseudophakia indicating that joint-representation learning with adaptive gating can capture interactions between different types of diagnostic cues. Importantly, prospective on-device testing demonstrated a nearly equivalent performance of 0.96 AUROC compared to offline evaluation, supporting feasibility for near-real-time, offline use in the field.

In clinical practice, ophthalmologists naturally combine visual examination cues with clinical context for decision-making. Age has been shown to be strongly connected to cataract severity due to both etiology and lack of access to care in LMICs.^10^ Visual acuity is a direct functional measure of the effect of lens opacity on vision and is used as surgical referral criteria.^38^ Pinhole acuity is used by eye care providers to distinguish cataract-related vision loss from other causes such as refractive error or retinal disorders.^39^ Thus, we hypothesized that a lightweight multimodal machine learning model trained on data from a large patient population may lead to significant improvements in lens classification. Our development replicates this clinical decision-making process. The tabular-only GBDT learned a hierarchy of features, while the late-fusion meta-learner prioritized the tabular probability of clear lens and the image probabilities of immature cataract and pseudophakia, demonstrating the idea that functional context could be more useful in identifying healthy cases whereas image-based cues could be more useful in identifying disease pathology. Early fusion further improved separation for immature and mature cataracts, likely because the learnable gate with average weights 0.808 and 0.907 for image and tabular data, respectively, allowed the network to rebalance information flow.

Recent anterior segment AI reports have shown multi-disease feasibility, also indicating potential for our pipeline to extend beyond cataract to broader anterior segment triage within the same capture workflow.^14^ Earlier cataract AI developments primarily focused on images from specialized cameras and without clinical context, limiting generalizability and field utility.^41^ Previously developed ophthalmic AI systems have achieved high accuracy in controlled settings but required substantial compute or advanced imaging systems.^42^ By advancing a multimodal, smartphone-deployed lens classification system capable of on-device deployment without loss of performance, our study overcomes accuracy and deployment challenges that have limited broad implementation.^41^ Future work will focus on multi-center evaluation, continuous and/or quantitative cataract grading (e.g., LOCS III scale), and extension to other anterior segment disorders. While models were developed with data from multiple hospitals, future external, prospective validation will include additional hospitals to further assess generalizability across pathologies and geographies. This will also enable assessment of model accuracy across a broader number of CHWs or technicians with varied training levels. Advanced image quality assessment could also be automatically performed and used as an input to encourage robustness to data quality. More sparsity in gating weights could allow more adaptive reasoning, which could better mirror clinical thought processes. Additional tabular features might further improve performance for lens classification and for triage of other anterior segment disorders, but may increase data collection burden or decrease generalizability.^43^

In summary, the multimodal early fusion model can classify lens status accurately using only a smartphone-based diffuse illumination image, patient age, and visual function measurements without performance loss when run on-device. Fusion architecture offers performance advantages over single modality classification, and more closely resembles clinical reasoning. This approach provides a feasible pathway to decentralized cataract screening and referral in low resource environments.

## Data Availability

All data produced in the present work are contained in the manuscript.

